# From Recognition to Reimbursement: An Assessment of State Medicaid Coverage Policies for Genetic Counselors and a Path Forward

**DOI:** 10.64898/2026.07.22.26358669

**Authors:** Philip D. Connors, Yue Guan, Cynthia A. James, Julian Polaris, Blair Cantfil, Colleen A. Campbell

## Abstract

**Purpose:** As genomics permeates all healthcare specialties, genetic counseling in conjunction with genetic testing is broadly recommended. Improving access to genetics professionals is crucial for Americans insured by Medicaid. The purpose of this study was to conduct a comprehensive review of Medicaid policies for genetic counseling performed by Certified Genetic Counselors (CGC®).

**Methods:** Fee-for-service Medicaid policies across 50 states and Washington DC were reviewed and coded. Four states with exemplary policies were identified, and CGC managers in two of these were surveyed regarding the real-world effects of these policies.

**Results:** As of 2024, 20 states (39%) had a published policy for genetic counseling with most (N=16, 80%) expressly covering genetic counseling in connection with any covered genetic test. Of the states without a policy, 12 (24%) mention genetic counseling in the context of scenario-specific policies, and 19 (37%) have no published policy. Twenty states explicitly cover CGC services, while 2 exclude CGCs as service providers. CGC managers in Indiana and Michigan confirmed the policies identified as exemplary successfully led to reimbursement of CGC services.

**Conclusion:** There is significant variability in Medicaid coverage for genetic counseling. Comprehensive policies are needed to support patient access to genetics professionals, including CGCs.

## INTRODUCTION

As genomic technologies continue to develop and impact the daily provision of healthcare, access to clinicians specifically trained in genetics, genomics, and personalized medicine is vital to meeting the needs of patients across the country. Both clinical genetics professional organizations - such as the American College of Medical Genetics and Genomics (ACMG) and the National Society of Genetic Counselors (NSGC) - and non-genetics clinical specialty societies develop practice guidelines and policy statements that outline the importance of access to genomics providers when genetic information may impact the provision of care for patients. As genetic testing and counseling permeate healthcare via primary care provision, chronic disease management, and other common care spaces, numerous organizations have highlighted the necessity of the clinical genomics workforce to their practice areas^1,2,3,4,5,6^. Virtually all guidelines that discuss access to testing for potentially heritable disease risk cite the importance of informed consent and thorough genetic counseling to ascertain the risks and benefits of utilizing genetic technologies to predict, explain, or prevent these conditions and to guide management. Access to these clinicians is paramount to providing safe and appropriate genomic healthcare, as the number of genetic tests registered with the NIH Genetic Testing Registry is now nearly 68,000^7^.

In the United States (US), genetic counselors have been key to addressing this growing need. The genetic counseling profession is one of healthcare’s newest clinician categories, having graduated its first Master’s-trained professionals in 1971. New Jersey was the first state to pass legislation to license genetic counselors in 1996^8^. Today, 39 of the 50 states and Washington DC do so, recognizing the growing recognition of the importance of ensuring appropriate training, credentials, and oversight for this critical workforce^9^. The National Coordinating Center for the Regional Genetics Networks (NCC) Medical Genetics Workforce Workgroup completed a workforce analysis and estimated that there were 9,293 genetics professionals in the US in 2023, and that a significant majority (71.5%) of those professionals are genetic counselors^10^. This segment of the workforce continues to expand rapidly - the National Society of Genetic Counselors 2024 Professional Status Survey identified a 100% growth in the profession from 2014 to 2024, and estimates a further 100% expansion of Certified Genetic Counselors (CGCs) by 2034 to total more than 11,000 clinicians in North America^11^. Still, many areas of the country have limited access to genetic counselors^12,13,14^.

Evidence suggests that genetic counselors ensure the appropriate utilization of genomic testing compared to when genetics professionals are not involved in care, saving both payors and patients money^15^.

While the genetic counseling profession continues to grow to meet expanding patient and provider needs, access to genetics clinicians remains limited for many patients, with coverage policies for genetic counseling largely dictating an individual’s access to these recommended standards of care. Furthermore, private payers are inconsistent in their policies regarding genetic counseling^16^. In addition, Medicare recognition of genetic counselors via Congressional legislation would bring meaningful access to genetics services for the 69 million Americans who are over the age of 65, permanently disabled, or living with end stage renal disease^17^. This study focuses on Medicaid, which covers more than 75 million low- and middle-income people, including almost one out of four children and adolescents^18^. Because each state defines its own coverage policies (subject to broad federal guardrails), states vary significantly in their policies on CGCs. NSGC cites increasing the number of states with Medicaid [genetic counselor] recognition and reimbursement policies for these services as a strategic priority in its 2025-2027 Strategic Plan^19^. Reys et al (2025) found that states with licensure are more likely to permit CGCs to enroll in state Medicaid programs and bill independently, and reviewed variation across states in terms of whether the Current Procedural Terminology (CPT®) code 96040 is listed in states’ Medicaid fee schedules and, if so, at what rate. That analysis acknowledged, however, fee schedules do not provide a full picture of state coverage requirements and limitations.

As with any service, coverage policies have a big impact on provider staffing and patient access. In addition, specific billing codes can help ensure accurate billing and reimbursement. NSGC and ACMG advocated for a new CPT® code for genetic counseling by genetic counselors (CPT® code 96040 was updated and replaced by 96041 effective January 1, 2025). In light of the new code and the 2025-2027 NSGC Strategic Plan objective to increase Medicaid coverage of genetic counselor services, the 2024 NSGC Board of Directors commissioned a comprehensive landscape assessment to systematically characterize Medicaid coverage policies and reimbursement for genetic counseling services provided by genetic counselors across the US, and to examine how these policies intersect with related coverage mechanisms, including biomarker legislation. We subsequently surveyed genetic counselor managers in states with the strongest genetic counseling Medicaid policies for feedback on the real-world application and implications of these policies.

This study offers a comprehensive review of state Medicaid coverage policies for genetic counseling and CGCs, validates the effectiveness of model state policies, and provides recommendations on how states (and other payers) can update and clarify their policies to promote access to high-value CGC services.

## METHODS

Between May and September 2024, a team of researchers at Manatt Health, supervised by an attorney (JP), systematically reviewed publicly available fee-for-service (FFS) Medicaid policies across all 50 states and Washington DC. The review encompassed each state’s Medicaid State

Plan, statutes, regulations, fee schedules, provider manuals, and other policy guidance. Managed care contracts were not included in the analysis.

This study focused on policies governing coverage and reimbursement for genetic testing and genetic counseling services. For each state, researchers coded information across four major domains (**Table 1**). The coding excluded coverage and billing policies specific to routine prenatal testing for pregnant individuals.

**Table 1.** Coding domains and guiding questions for the Medicaid FFS policy review.

| Domain | Description / Guiding Questions | Example Coding Elements |
| --- | --- | --- |
| **1. Coverage Policy for Genetic Testing** | Presence, scope, and specificity of written coverage policy for genetic testing. | (a) Comprehensive biomarker law that applies to Medicaid FFS (based on model language from the National Council of Insurance Legislators or the American Cancer Society Cancer Action Network); (b) A Medicaid policy defining coverage criteria for genetic tests in general; (c) A Medicaid policy defining coverage criteria for specific tests or conditions only. |
| **2. Coverage of Genetic Counseling Services** | Presence, scope, and specificity of coverage policy for genetic counseling. | Whether coverage is limited to, or required in, specific clinical scenarios. |
| **3. Coverage of Services by Certified Genetic Counselors** | Recognition and billing status of genetic counselors as providers. | (a) Whether genetic counselors are explicitly named or covered by implication (e.g., “all qualified providers” or “per NCCN guidelines”); (b) Whether they can bill independently or whether their services are billed by a supervising physician. |
| **4. Reimbursement for Certified Genetic Counselor Services** | Billing codes used for reimbursement. | CPT® 96040 (30-min increments for genetic counselor services) (*note: this code has now been superseded by 96041*); HCPCS S0265 (15-min increments under physician supervision). |

In September 2025, authors PFC, YG, CAJ, and CAC - 2024 NSGC Board leaders with clinical, research, and policy expertise - reviewed data and selected two states (Indiana and Michigan) as models for Medicaid reimbursement policies for genetic counseling. An online survey was developed to explore whether the policies that appeared to be exemplary models for genetic counseling reimbursement worked well in practice within the selected states. Survey domains included policy awareness, billing and reimbursement practices, institutional implementation barriers and facilitators, and perceived impact on access to services. Using the NSGC member database, a convenience sample (n=4) was identified consisting of two genetic counselor managers in each state: one working at an academic medical center and the other in a community hospital setting. Targeted outreach was conducted in February - March 2026 to these individuals.

A qualitative, descriptive analysis of CGC managers’ survey responses to examine how stated policy intent aligned with real-world effects on access to and reimbursement for genetic counseling services. This survey study was determined to be exempt by the University of Iowa Institutional Review Board (University of Iowa IRB #202511444). A copy of the questionnaire is available in the Supplemental Material.

## RESULTS

### Coverage of Genetic Counseling Services in General

Based on this deep review of the Medicaid FFS coverage policies of all 50 states and the District of Columbia, 20 states (39%) have a published general policy describing coverage for genetic counseling as of 2024, outlined in **Figure 1**. Among these 20 states, most (N=16, 80%) expressly cover genetic counseling in connection with any covered genetic test, including 13 that *require* genetic counseling as a condition of coverage for some or all genetic tests.

**Figure 1.**
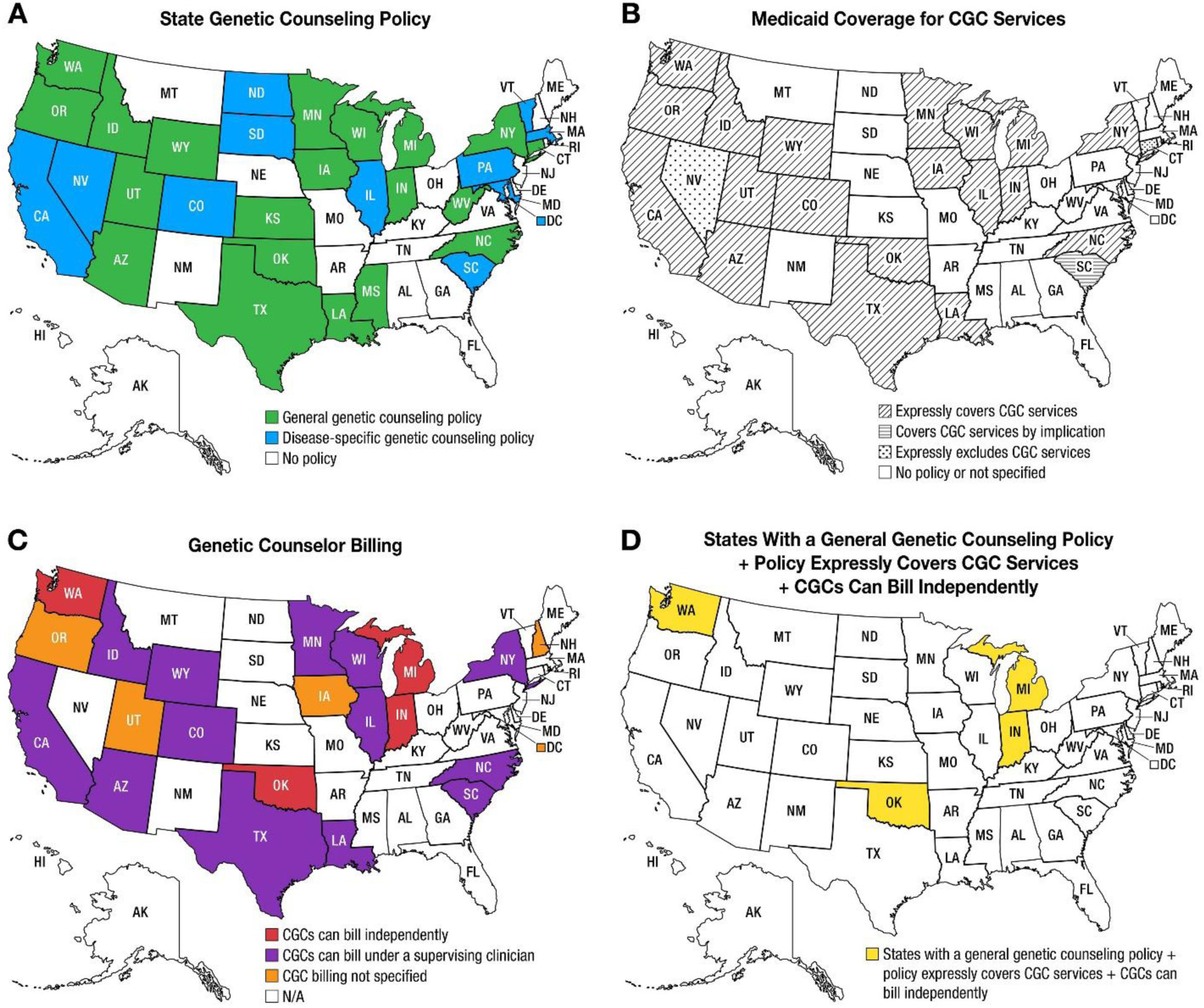
Medicaid Coverage Policy for Genetic Counseling and CGC Services. **Map A** identifies states with Medicaid FFS general coverage policies for genetic counseling, disease-specific coverage policies for genetic counseling, and states without policies. **Map B** identifies if these policies expressly cover CGC services, cover CGC services by implication, or exclude CGC services from coverage. **Map C** identifies if these policies allow for independent billing of CGC services, billing under a supervising clinician, or is not otherwise specified. **Map D** identifies the 4 states where there is 1) a general FFS policy, 2) CGC services are expressly covered, and 3) CGCs can bill independently for their services.

**Table 2** summarizes the results of the analysis, categorizing genetic counseling coverage policies, coverage of services by genetic counselors, and billing policies, including whether CGCs can bill independently and whether 96040 is included in the fee schedule for each state. Figure 1 provides a visual representation of this information in a US map. As can be appreciated, there is considerable heterogeneity in each domain, resulting in quite different Medicaid practice environments for genetic counselors. Among the remaining 31 programs without a general policy, 12 (24%) mention genetic counseling in the context of disease or scenario-specific policies, such as for hereditary breast and ovarian cancer or genetic testing during pregnancy. Of these, 6 states (12%) require genetic counseling as a condition of coverage for at least some genetic tests. The remaining 19 states (37%) have no published policy regarding genetic counseling.

**Table 2.** Summary of state Medicaid policies regarding genetic counseling. FFS= Fee-for-Service; CGC® = Certified Genetic Counselor.

| State | Does the state have a coverage policy for genetic counseling? | Coverage of Services by CGCs? | CGCs bill independently? | CPT® Code 96040 listed in fee schedule |
| --- | --- | --- | --- | --- |
| AK | No | Not specified | N/A | No |
| AL | No | Not specified | N/A | No |
| AR | No | Not specified | N/A | No |
| AZ | Yes, General Coverage Policy | Yes, Expressly | Under Supervising Clinician | No |
| CA | Yes, only in specific clinical scenarios | Yes, Expressly | Under Supervising Clinician | No |
| CO | Yes, only in specific clinical scenarios | Yes, Expressly | Under Supervising Clinician | Yes |
| CT | Yes, General Coverage Policy | No | N/A | N/A |
| DC | Yes, only in specific clinical scenarios | Not specified | Not specified | Yes |
| DE | No | Not specified | N/A | No |
| FL | No | Not specified | N/A | No |
| GA | No | Not specified | N/A | No |
| HI | No | Not specified | N/A | No |
| IA | Yes, General Coverage Policy | Yes, Expressly | Not specified | Yes |
| ID | Yes, General Coverage Policy | Yes, Expressly | Under Supervising Clinician | Yes |
| IL | Yes, only in specific clinical scenarios | Yes, Expressly | Under Supervising Clinician | Yes |
| IN | Yes, General Coverage Policy | Yes, Expressly | Independent | Yes |
| KS | Yes, General Coverage Policy | Not specified | N/A | No |
| KY | No | Not specified | N/A | No |
| LA | Yes, General Coverage Policy | Yes, Expressly | Under Supervising Clinician | Yes |
| MA | Yes, only in specific clinical scenarios | Not specified | N/A | Yes, but only for youth <21 |
| MD | Yes, only in specific clinical scenarios | Not specified | N/A | No |
| ME | No | Not specified | N/A | No |
| MI | Yes, General Coverage Policy | Yes, Expressly | Independent | Yes |
| MN | Yes, General Coverage Policy | Yes, Expressly | Under Supervising Clinician | Yes |
| MO | No | Not specified | N/A | No |
| MS | Yes, General Coverage Policy | Not specified | N/A | No |
| MT | No | Not specified | N/A | No |
| NC | Yes, General Coverage Policy | Yes, Expressly | Under Supervising Clinician | Yes |
| ND | Yes, only in specific clinical scenarios | Not specified | N/A | Yes |
| NE | No | Not specified | N/A | No |
| NH | No | Not specified | Not specified | No |
| NJ | No | Not specified | N/A | Yes |
| NM | No | Not specified | N/A | No |
| NV | Yes, only in specific clinical scenarios | No | N/A | N/A |
| NY | Yes, General Coverage Policy | Yes, Expressly | Under Supervising Clinician | Yes |
| OH | No | Not specified | N/A | N/A |
| OK | Yes, General Coverage Policy | Yes, Expressly | Independent | Yes |
| OR | Yes, General Coverage Policy | Yes, Expressly | Not specified | Yes |
| PA | Yes, only in specific clinical scenarios | Not specified | N/A | Yes |
| RI | No | Not specified | N/A | No |
| SC | Yes, only in specific clinical scenarios | Yes, by Implication | Under Supervising Clinician | Yes |
| SD | Yes, only in specific clinical scenarios | Not specified | N/A | No |
| TN | No | Not specified | N/A | N/A |
| TX | Yes, General Coverage Policy | Yes, Expressly | Under Supervising Clinician | Yes |
| UT | Yes, General Coverage Policy | Yes, Expressly | Not specified | Yes |
| VA | No | Not specified | N/A | No |
| VT | Yes, only in specific clinical scenarios | Not specified | N/A | No |
| WA | Yes, General Coverage Policy | Yes, Expressly | Independent | Yes |
| WI | Yes, General Coverage Policy | Yes, Expressly | Under Supervising Clinician | No |
| WV | Yes, General Coverage Policy | Not specified | N/A | No |
| WY | Yes, General Coverage Policy | Yes, Expressly | Under Supervising Clinician | Yes |

### Coverage of Services by Certified Genetic Counselors

Of the 32 programs that have either a general coverage policy or disease-specific coverage policy for genetic counseling services, 20 expressly cover genetic counseling *by genetic counselors*, including 17 states that reimburse using CPT® 96040.

Of the remaining 12 states with general or disease-specific coverage for genetic counseling services, 2 expressly exclude genetic counselors as service providers, and the remaining 10 do not specify a provider type. These nuances are available for review in Supplementary Figure 1.

A total of 4 states (Indiana, Michigan, Oklahoma, and Washington) were identified with policies that expressly covered genetic counseling services, allowed genetic counselors to bill independently, and listed CPT® code 96040 in the fee schedule. Of these, Michigan and Indiana were selected as states meeting these criteria and most appropriate to analyze as case studies due to: 1) Representativeness - These two state programs balance both urban and rural healthcare settings in their coverage system, with similar overall population density and Medicaid expansion under the Affordable Care Act; and 2) Generalizability - While over the past decade Michigan has been primarily led by Democratic leadership in their state house and legislature, Indiana has been primarily led by Republican leadership. Both states’ average annual household incomes are slightly below the national median when considering the cost of policy implementation from state and federal tax revenue. Of note, there are other states that enroll CGCs for Medicaid program reimbursement - however, these states are reimbursing for these services in the absence of a clearly defined or publicly available policy, and therefore were not selected for further analysis.

### Indiana Case Study

Indiana requires genetic counseling as a condition of coverage for genetic tests. While acknowledging that genetic counseling can be performed by a range of provider types, Medicaid agency guidance confirms that “providers cannot use the title ‘genetic counselor’ unless licensed as such,” and that genetic counselors can enroll for the specific and sole purpose of furnishing genetic counseling services.^20^

The two surveyed Indiana CGC managers confirmed that the providers at their institutions were credentialed with Medicaid in 2024, billed independently for their services and were reimbursed by Medicaid. Both participants described Indiana’s policy as “effective”, and one stated it is effective because it allows CGCs to be *“reimbursed for care and our licensure allows testing to be ordered in GC’s name--both improve access to care and permit GCs to work at top of scope. Such policies improve our ability to add new positions,”* although the respondent expressed a concern about low reimbursement rates.

Respondents emphasized the advocacy value of physician champions “who knew who genetic counselors were and our value,” including a primary care provider who relied on the skillset of genetic counselors and was now working on Medicaid policy in the state.

Other considerations when developing a policy reflected on opportunities to improve patient access to care:

> *“Our state’s Medicaid policy requires a referral to be issued from a Medicaid-enrolled [physician, physician assistant, or nurse practitioner] that specifically requests genetic counseling in order for [genetic counselor] services to be reimbursed. While I do think this policy helps maintain communication between patients, general healthcare providers and the genetics team, there have been instances where this has put undue pressure on clinical workflows to get referrals and has resulted in some delays in scheduling for patients wishing to self-refer or for those who do not have an established [primary care physician] or other provider willing to provide referral.”*

### Michigan Case Study

The Michigan Medicaid agency provides a clear description of when genetic counseling is covered by genetic counselors and other practitioner types: Genetic counseling is “covered when provided in consideration of, or in conjunction with, genetic testing, or provided in relation to a genetic or congenital condition. Services are considered medically necessary when there is an expectation that a genetically inherited or acquired condition exists, and the beneficiary displays clinical features or is at risk of inheriting the disease/condition based upon factors including, but not limited to, personal history, family history, documentation of a genetic mutation, and/or ethnic background." [Provider Manual, s.5.3]^21^

The two surveyed Michigan CGC managers confirmed that CGCs at their institution were credentialed with Medicaid in 2024, billed Medicaid independently for their services, and were reimbursed by Medicaid. One respondent described Michigan’s policy as “effective,” including *“clear messaging from the State about the policy to cover 96040.”* As background, they noted that the code’s introduction supported advocacy efforts to permit genetic counselors to bill independently.

## DISCUSSION

The results demonstrate a wide spectrum of policies regarding genetic counseling services. We posit that a general coverage policy for genetic counseling by genetic counselors is the gold standard for ensuring clarity and transparency in providing access to CGC services in state Medicaid programs and other payors across the country. These data suggest there is significant lack of clarity for the majority of states regarding where CGC services can be successfully provided and reimbursed, as it is not explicitly clear to providers and health care revenue cycle management teams how billing should be incurred and when appeals of denied reimbursement are warranted. It is especially confusing in the instances where genetic testing policies are implicitly instructive in covering CGC services, but CGC enrollment and coverage lack explicit guidance or support.

As genomic medicine and genetic testing are integrated throughout healthcare and a growing number of practice guidelines recommend genetic counseling provided by genetics professionals, healthcare institutions seek to grow their genetics workforce to meet patient and provider needs. However, limited reimbursement of genetic counseling services is a barrier to growing such service delivery models. Academic medical centers have historically absorbed costs of unreimbursed care or offset costs with research grants, enabling genetic counselors to serve as physician extenders - increasing physician work volume and efficiencies, but limiting overall clinical throughput of patients seeking genetics services. This is unlikely to be a sustainable model for provision of genomics services. Additionally, community hospitals may not have the same resources or ability to hire genetic counselors without appropriate reimbursement for their services, further limiting access for patients to receive genetic counseling services - especially those in rural or underserved areas.

Regardless of setting, physicians and administrators who want to include genetic counselors on their team are key advocates to improve policies, as specifically noted by survey respondents. This advocacy is critical to generate a revenue stream to help offset clinic personnel costs. To address the growing need for genetic counseling services, advocacy by physicians, advanced practice providers, health system and hospital administrators, patient advocates, and professional societies for improved reimbursement of genetic counselor services is ongoing and essential.

Our findings both build upon and extend prior work, demonstrating greater complexity in state Medicaid coverage policies for genetic counseling than previously documented. Reys et al cited that 26 of 51 (51%) state Medicaid plans have public-facing policies that suggest reimbursement for genetic counseling – one of which permits reimbursement only when provided under physician supervision. They note that while 34 states had implemented genetic counseling licensure legislation in 2024, only 11 enrolled genetic counselors as providers with their Medicaid plans^8^. It is important to note, licensure does not equate to genetic counseling payor policies nor automatically result in reimbursement for genetic counselor services. Our current study demonstrates the importance of distinguishing enrollment from potential coverage. For example, Connecticut and Nevada have genetic counseling coverage policies but do not cover services provided by genetic counselors. These data also highlight the disconnect between the publication of CPT® codes in a fee schedule and coverage policies: in New Jersey, there is *not* a genetic counseling coverage policy, yet CPT® 96040 is listed in the fee schedule, whereas Arizona has a general genetic counseling coverage policy and CPT® code 96040 is *not* listed fee schedule. Our work also identified one state that lists CPT® 96040 but has no coverage policy to refer to, and a number of states with genetic counseling coverage policies that suggest genetic counseling by genetic counselors is billable but does not list a CPT® code to recoup for that service.

Our study adds that publicly available fee schedules do not entirely explain the complexity of genetic counseling reimbursement policies. For example, Reys et al found 30 states listed 96040 in the fee schedule, whereas the current study found 32 states had some kind of policy for genetic counseling. However, having a policy does not mean the CPT® code 96040 is in the fee schedule, and in fact, the situation is much more nuanced across the country with 21 states having some kind of genetic counseling policy and 96040 listed in the fee schedule; however, who can bill that code differs by state, demonstrating extreme variability nationwide, Supplementary Figure 1. Moreover, many state fee schedules contain caveats that a code being listed in the fee schedule does not necessarily mean the code is covered.

Certified genetic counselors at academic medical centers and community hospital settings in Indiana and Michigan reported receiving reimbursement for their genetic counseling services, suggesting a clearly written Medicaid policy can benefit access to genetic counseling services regardless of clinical setting. As more states adopt policies that expressly cover genetic counselor services, further studies are warranted to explore differences in experience, reimbursement, and the ability to hire genetic counselors in diverse clinical settings.

As evidenced by this work and previous efforts regarding cell and gene therapy access^22^, successful payer policies that can be implemented consistently and effectively across different types of institutions must contain the following elements: 1) the service to be provided, 2) coverage conditions, 3) the type of provider that can provide the service, 4) how the provider can bill the service, and 5) the code to be listed in the fee schedule. Without all these elements, ambiguities or unintentional limitations can lead to ineffective policy. Furthermore, extraneous requirements such as requiring referrals from specific provider types for CGC services to be reimbursed can diminish the policy’s efficacy in achieving the goal of providing patients access to services. When the distribution of genetics providers tends to be greater in metropolitan areas and cities with academic medical centers, such policy riders inadvertently maintain access barriers for individuals who live in rural or underserved areas.

Without clear coverage policies, optimizing service delivery and studying its impact on quality and economic measures for Medicaid populations is challenging. Concurrent with lack of coverage under Medicare, there are no large studies of CGC impact on quality, safety, and efficiency in publicly insured cohorts in the United States. Available data in other populations and disease-specific cohorts suggest heterogeneous patient impact^23, 24,25,26^, while the clinical necessity of the service remains undisputed and noncontroversial. Genetic testing is now a routine part of medical care, with test volume and spending rising rapidly^27^. Increasingly, studies show that a significant proportion of genetic tests are misordered relative to clinical guidelines, leading to unnecessary lab spend, specialist visits, and interventions^28^. Policies that recognize and enable certified genetic counselors providing genetic counseling will assist health plans in managing the growing demand for genetic testing and ensure evidence-based testing and access to care^15,29^. Individuals and organizations advocating for genetic counselor reimbursement may benefit from citing this growing body of literature in challenging economic environments, as seen in the context of disease-specific advocacy for hereditary breast and ovarian cancer syndrome genetic counseling and testing coverage^26^.

### Policy Advocacy Strategies

The results of this study informed the 2025-2027 NSGC Strategic Plan’s investment of resources to improve Medicaid recognition and reimbursement. These efforts are ongoing, and no single approach to increasing the number of high-quality policies is best. Some states may require legislation to add covered services, while others may be able to add new providers/services via regulatory means. Providers across the country must continue to assess their own local coverage status and engage their institutional resources to best understand the facilitators and barriers to access to CGCs for patient cohorts covered by Medicaid. As a public insurance product for those marginalized by socioeconomic status and other social determinants of health, genetic counseling represents a scalable, patient-centered care service focused on the most personal aspect of one’s health - genomic predictors of disease. Given the unique nature of these >50 state and district-administered programs, no one organization or program can solve these issues alone. Professional organizations must continue to work together, developing resources and relationships to support and empower clinician advocacy for appropriate reimbursement.

This study has several limitations. The policy review was restricted to Medicaid FFS policies and did not include analysis of managed care plan contracts or plan-specific coverage policies, which may differ from FFS in states that have adopted managed care. Additionally, the qualitative survey was limited to a convenience sample of four genetic counselor managers in two states, and findings should be interpreted with caution and may not be generalizable to other states or practice settings. The cross-sectional nature of the policy review reflects coverage policies as of 2024 and may not capture subsequent policy changes. Despite these limitations, this study provides a comprehensive and systematic assessment of state Medicaid coverage policies for genetic counseling services and offers actionable recommendations for policy improvement.

## CONCLUSION

Medicaid coverage of genetic counseling provided by genetic counselors is highly variable across the country and more nuanced than publicly available data. This study underscores the importance of clearly written policies in improving patient access to genetics professionals and genetic testing. When advocating for a clear policy, we recommend that states include the following elements: 1) a general coverage policy, agnostic to specific disease or condition, 2) genetic counseling services provided by genetic counselors are expressly stated, 3) genetic counselors are able to bill independently, and 4) the genetic counselor CPT® code, 96041, is listed in the fee schedule. Michigan and Indiana demonstrate that such policies can support independent billing, expand the genetics workforce, and improve patient access across both academic and community settings, and serve as potential models for other states. Clear policy can aid the state, employers, and providers in implementing the policy accurately, thereby reducing unnecessary costs while improving patient access to genetic services. Advocacy at the state level by multiple stakeholders, including physicians, other healthcare providers, patient advocacy organizations, and employers, is essential to advancing this goal.

## DATA AVAILABILITY

Data from this study are not publicly available.

## ACKNOWLEDGEMENTS

We would like to thank the 2024 National Society of Genetic Counselors Board of Directors: Colleen A. Campbell, Sara Pirzadeh-Miller, Carla McGruder, Salma Nassef, Deepti Babu, Austin Bland, Philip D. Connors, Cynthia A. James, Carrie Haverty, Chris Tan, Sheetal Parmar, Yue (Guan) Guan for their approval and funding of the landscape assessment.

We would like to thank Meghan Carey, Carrie Haverty, and Sara Pirzadeh-Miller for their review of the manuscript.

## AUTHOR CONTRIBUTIONS

Conceptualization: P.D.C., Y.G., C.A.J., C.A.C.; Data curation: J.P.; Methodology: P.D.C., Y.G., C.A.J., J.P., B.C., C.A.C.; Formal analysis: P.D.C., J.P., B.C., C.A.C.; Project administration: C.A.C.; Writing-original draft: P.D.C., C.A.C.; Writing-review & editing: P.D.C., Y.G., C.A.J., J.P., B.C., C.A.C.

## ETHICS DECLARATION

The survey study was determined to be exempt by the University of Iowa Institutional Review Board (University of Iowa IRB #202511444).

---

**Supplementary Figure 1.**
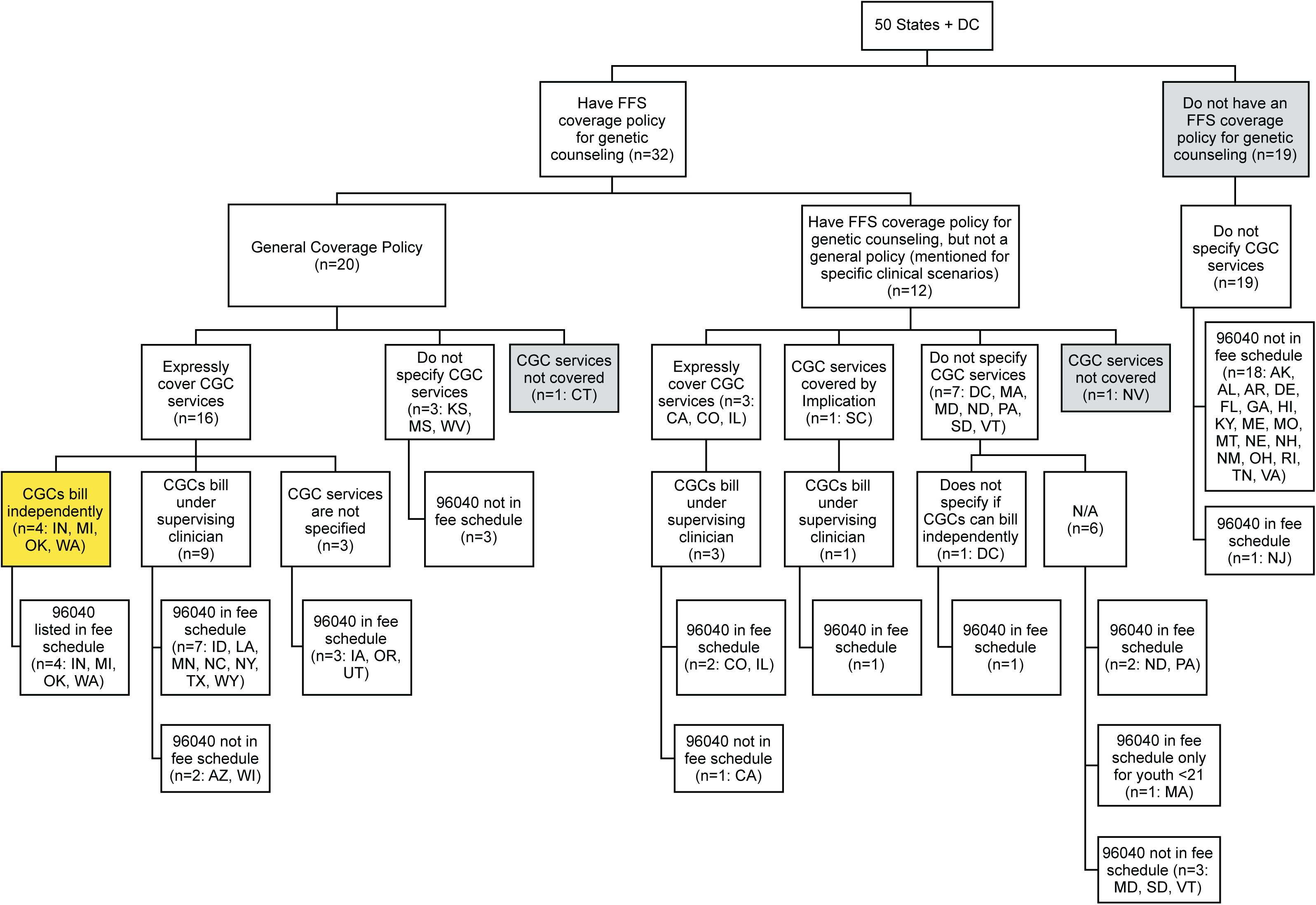
Summary of state Medicaid policies regarding genetic counseling. N = number of states that meet the criteria; CGC = Certified Genetic Counselor; Gold = states that have general genetic counseling policies, expressly cover genetic counseling services, allow genetic counselors to bill independently, and list CPT^®^ 96040 in the fee schedule; Gray = states that do not cover genetic counseling services provided by certified genetic counselors.

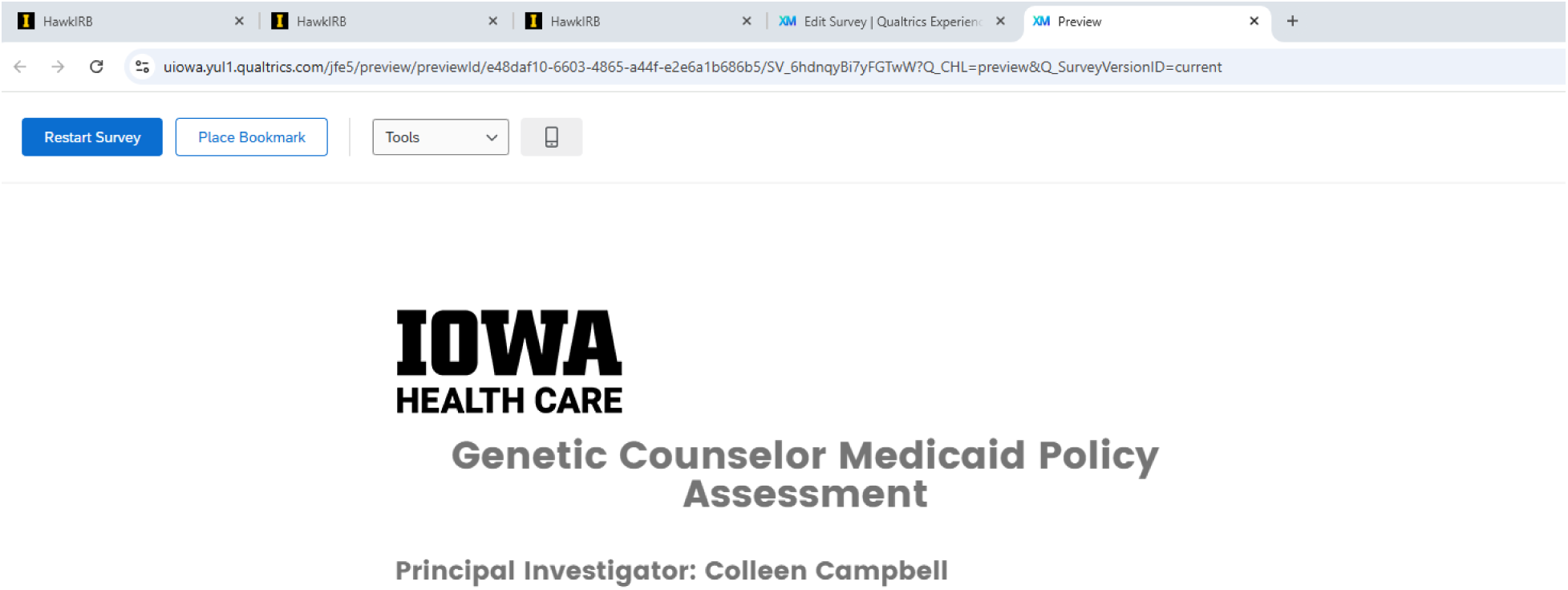
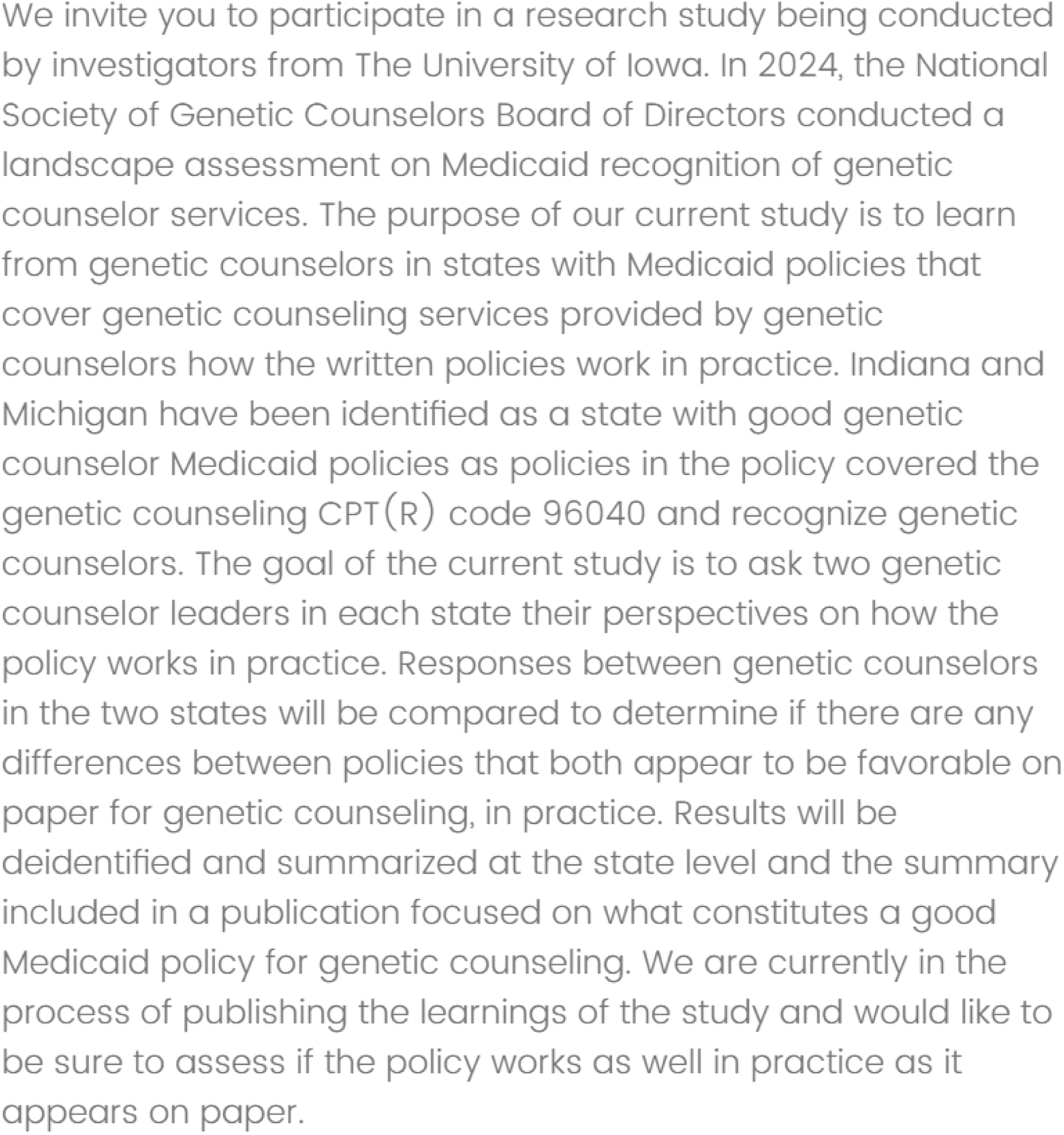
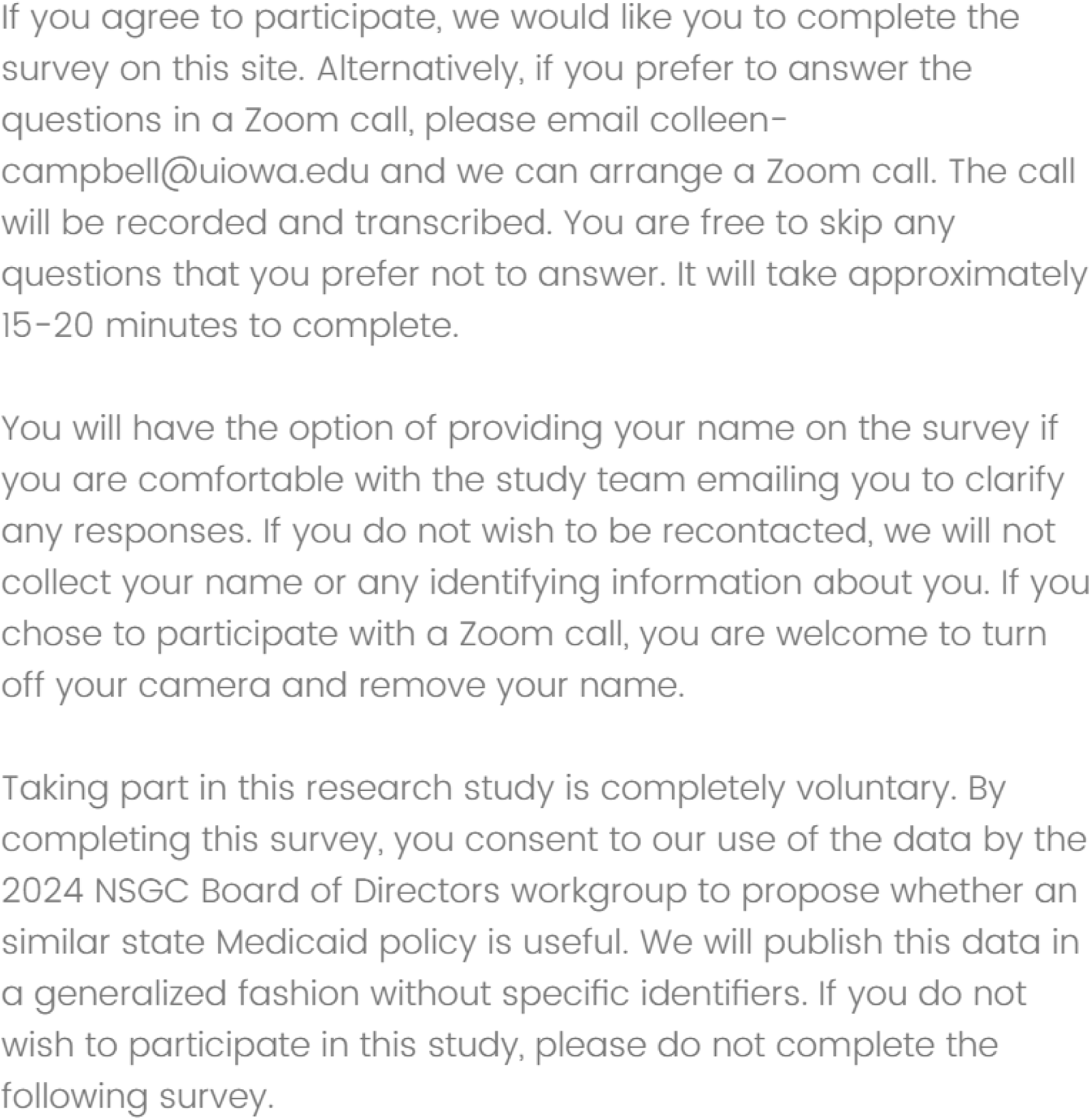
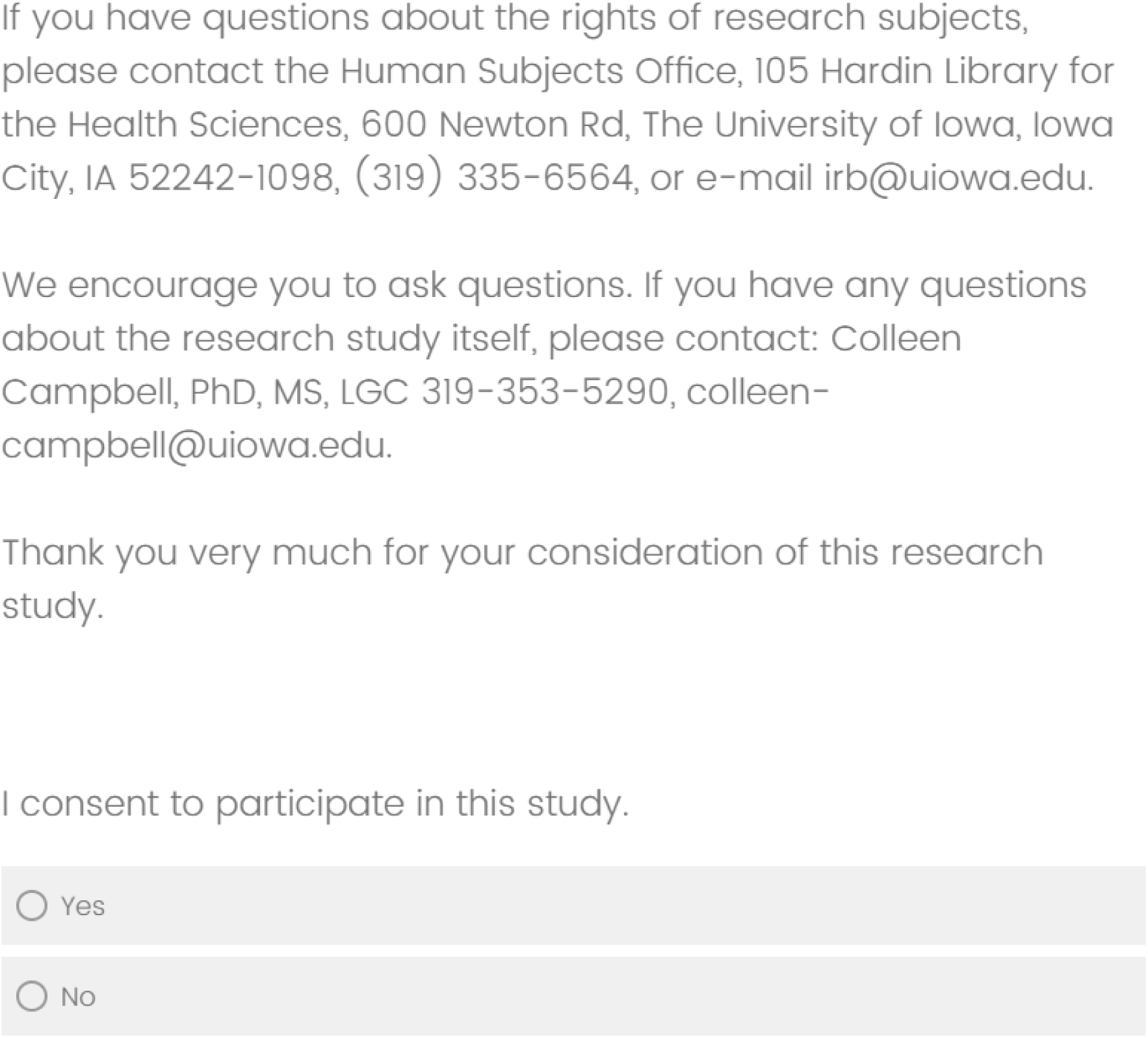
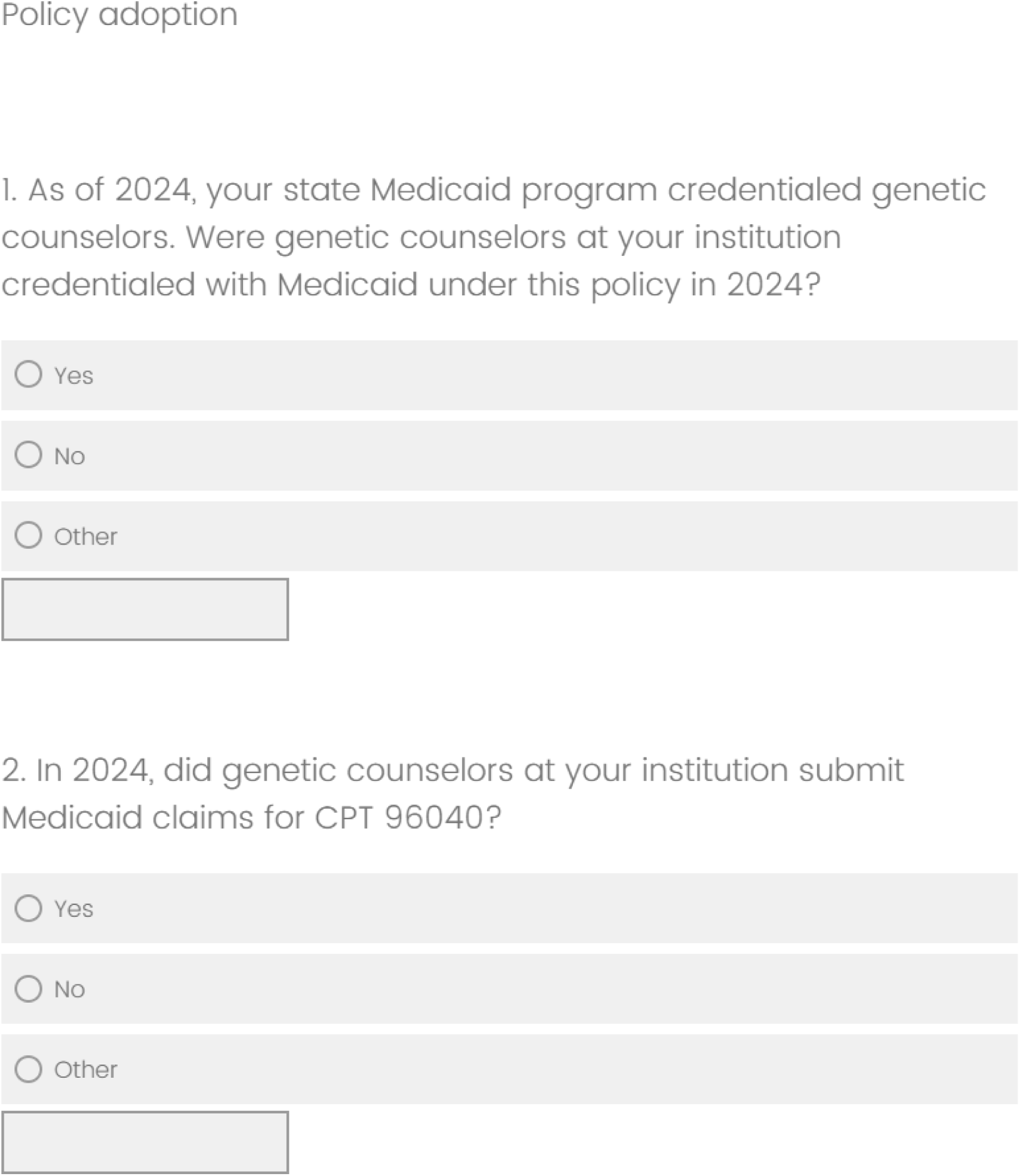
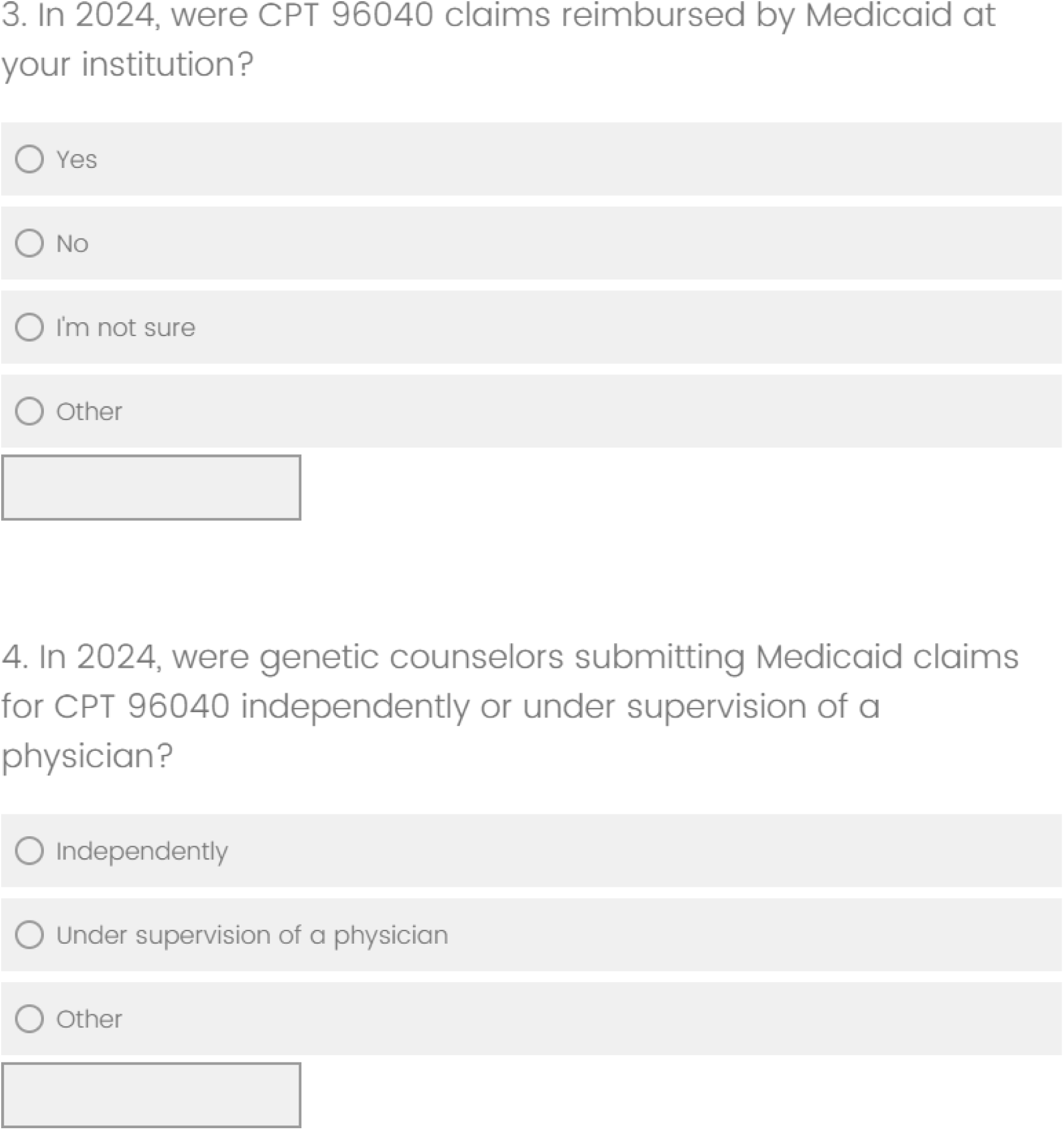
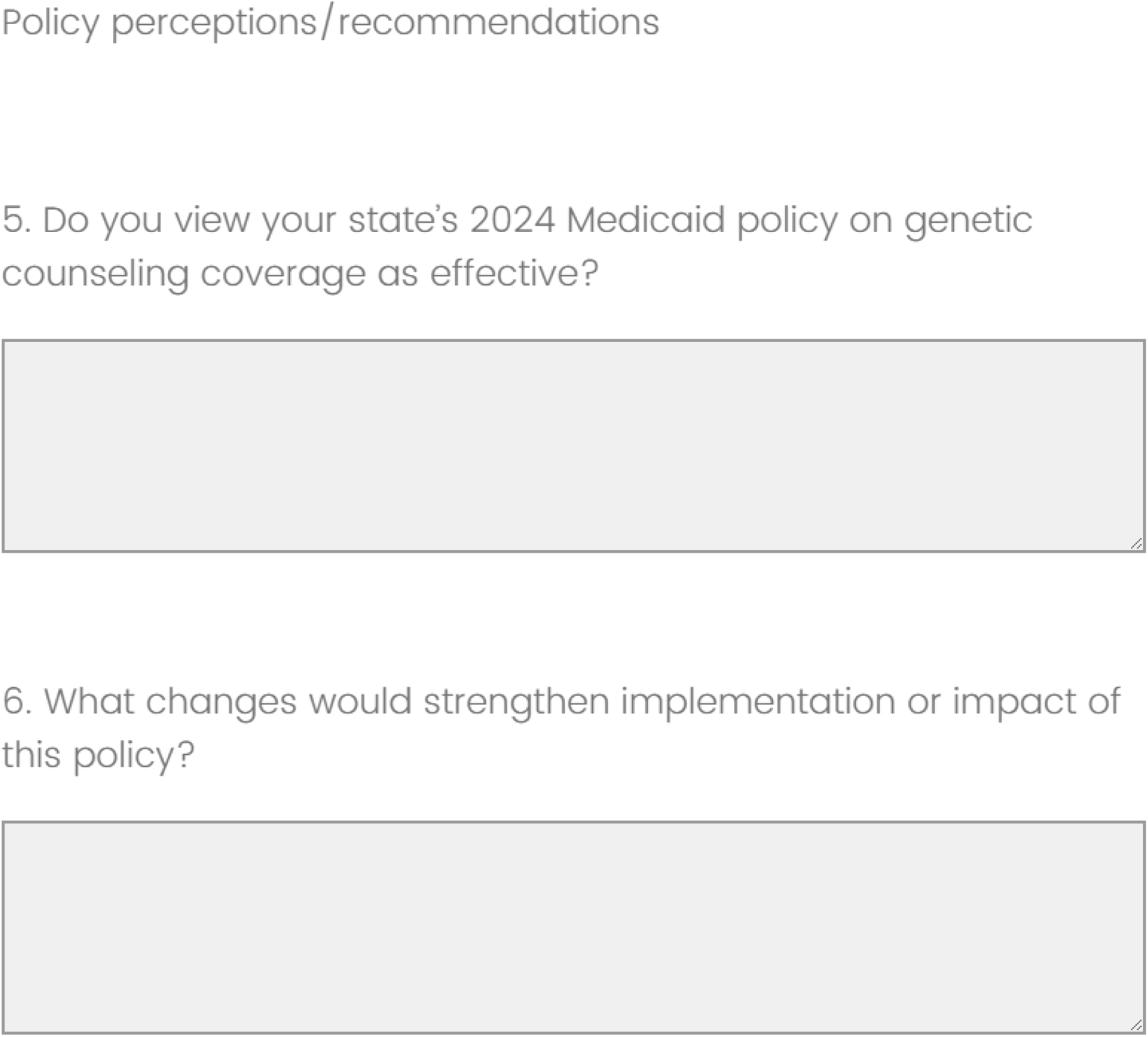
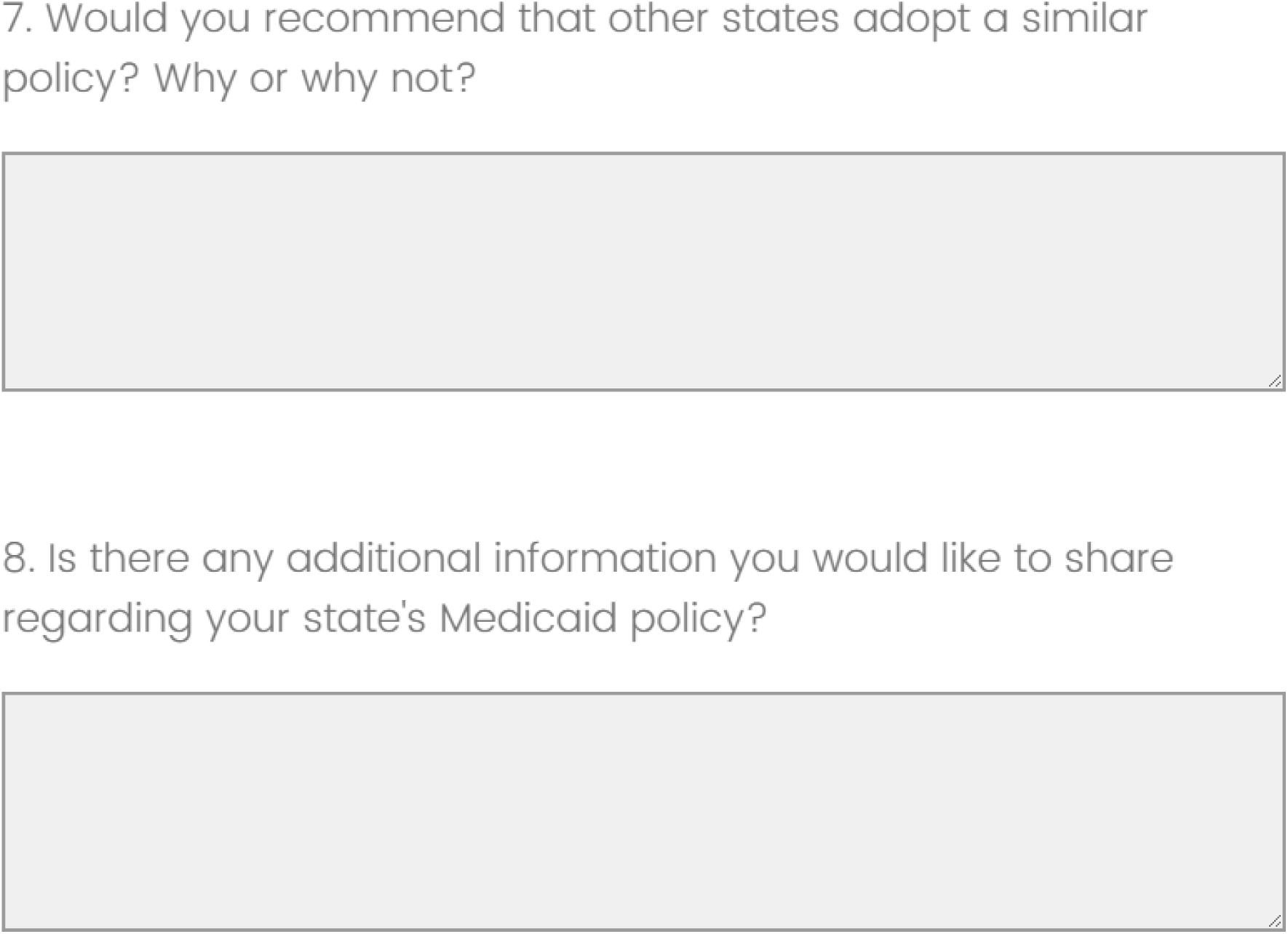
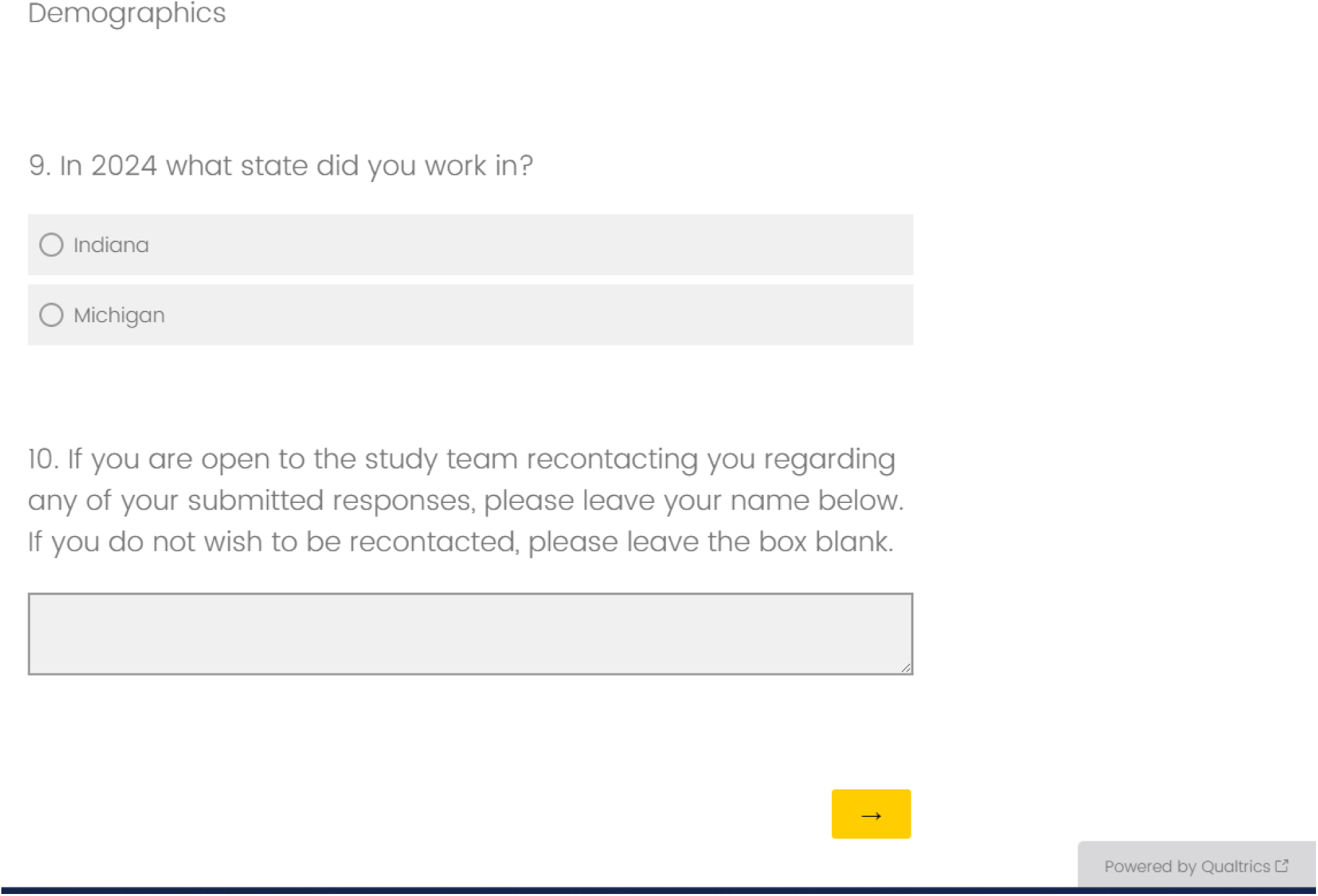

